# Machine-learning approach for Type 2 Diabetes diagnosis and prognosis models over heterogeneous feature spaces

**DOI:** 10.1101/2023.07.26.23293097

**Authors:** J.Ramón Navarro-Cerdán, Pedro Pons-Suñer, Laura Arnal, Joaquim Arlandis, Rafael Llobet, Juan-Carlos Pérez-Cortés, Francisco Lara-Hernández, Luis Alvarez, Ana-Bárbara García-García, Felipe Javier Chaves

## Abstract

This research aims to evaluate the Type 2 Diabetes (T2D) diagnosis and prognosis power from heterogeneous environmental, lifestyle and biochemistry data. Model estimation has previously addressed three main actions as: 1) Missing-value imputation using specific univariant and multivariant imputers accommodated to each particular feature; 2) Quasi-constancy detection in variables; 3) Constructing geographical pollution and rent data from municipality information. Next, different T2D diagnosis and prognosis models are fitted and evaluated, showing increasing performance as more specific features become available while the prediction cost rises as a consequence of requiring more specific data. Finally, four models are obtained: two of them for T2D diagnosis and the other two for T2D prognosis respectively, with performances ranging from 73.3 to 95.41 AUC-ROC. One pair of diagnosis and prognosis models were thought for a global testing that can be done in general locations by only asking general lifestyle-related questions. On the other hand, the other pair, which achieves higher performances, is thought to be applied in a clinical environment where it is easy to obtain more specific biochemistry measures.

## 1. Introduction

In the last 30 years, the prevalence of Type 2 Diabetes Mellitus (T2D) has increased significantly in adolescents and young adults, rising from an incidence of 117.07 per 100,000 population in 1990 to 183.36 in 2019 [1]. At ages under 30, women have higher mortality and disability-adjusted life years (DAILY) rates than men. This difference is reversed when we study a population aged over 30, except in countries with a low sociodemographic index. Countries with a low-medium and medium sociodemographic index have the highest age-standardized incidence rate. Studies [1] and [2] indicate that, in all regions, the leading risk factor for DAILY rate is the body mass index, although the proportional impact of other factors varies across regions and sociodemographic index. Other relevant factors are air pollution and smoking [3], household air pollution from solid fuels, and diets with low fruit intake, particularly in countries with a low sociodemographic index and high stress [4]. Early prevalence of T2D increases health problems as the population becomes older. As T2D appears earlier, the hyperglycemia duration is higher, and the pathological process speeds up, which drives towards a worsened glycemic control. The work of [5] shows evident gender inequality in general risk cardiopathies and T2D. Thus, from an economic perspective, T2D prevention produces excellent savings to health systems worldwide [6]. A better understanding of this disease would allow for a more accurate AI-based risk prevention models, and help to make patient personalized decisions to delay T2D appearance, which would result in an improved health status and longer lifetime for individuals, as well as significant economic savings for healthcare administrations.

T2D is a complex multifactorial disease that often requires complex models to achieve the best performances when attempting to predict it or its risk of appearance. In this work, several techniques and heuristics have been developed for relevant environmental factor identification, by increasing the quality of the original data and obtaining their structural relations. Thus, we challenged a double objective: 1) finding factors directly associated with T2D; and 2) obtaining T2D models for diagnosis and prognosis. The data utilised in this work, retrieved from T2D patients and individuals that develop the disease in the follow-up, includes heterogeneous clinical and environmental variables. In future work, we hope to use the developed heuristics and tools to include different kinds of genetic variations and individual information, in order to estimate prospective T2D detection models. These predictions are hoped to help preventing or delaying T2D appearance by proposing healthy lifestyles and scheduling temporary patient reviews to the correspondent risk group.

## 2. Material and methods

### 2.1. Data preparation and extension

The utilized dataset consists of 242 environmental features gathering basal information from 4617 anonymized patients from the Study, a populationbased study that has been described in detail [7]. All participants signed the informed consent for the Study. Valencian Clinical Hospital Ethics Committee aproved the work (references 2017.184 and 2031/036).

Features were grouped by their subject. Table 1 shows the number of variables for each type.

**Table 1:**
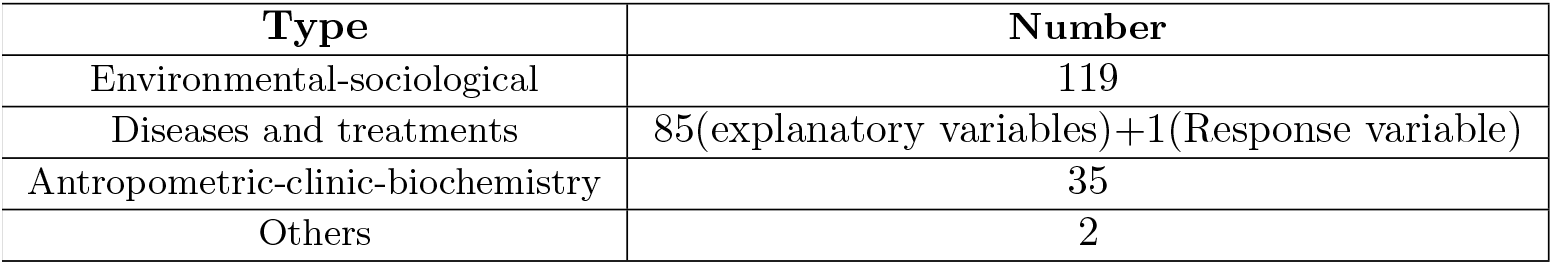
Number of features per thematic cluster.

| Type | Number |
| --- | --- |
| Environmental-sociological | 119 |
| Diseases and treatments | 85(explanatory variables)+1(Response variable) |
| Antropometric-clinic-biochemistry | 35 |
| Others | 2 |

The raw data exhibits several problems like missing values, quasy-constancy, string errors, etc, that severely affect its quality. Different processing steps must be applied to improve data quality and homogenization before further analysis is done. This includes feature filtering, string correction, missing data imputation, and quasi-constant features filtering. First, columns with over 90% of missing values are deleted. Then, string correcting is carried out using a weighted finite states transducer composition technique [8].

Figure 1 represents the whole processes involved in data preparation, including geospatial data construction from [9].

**Figure 1:**
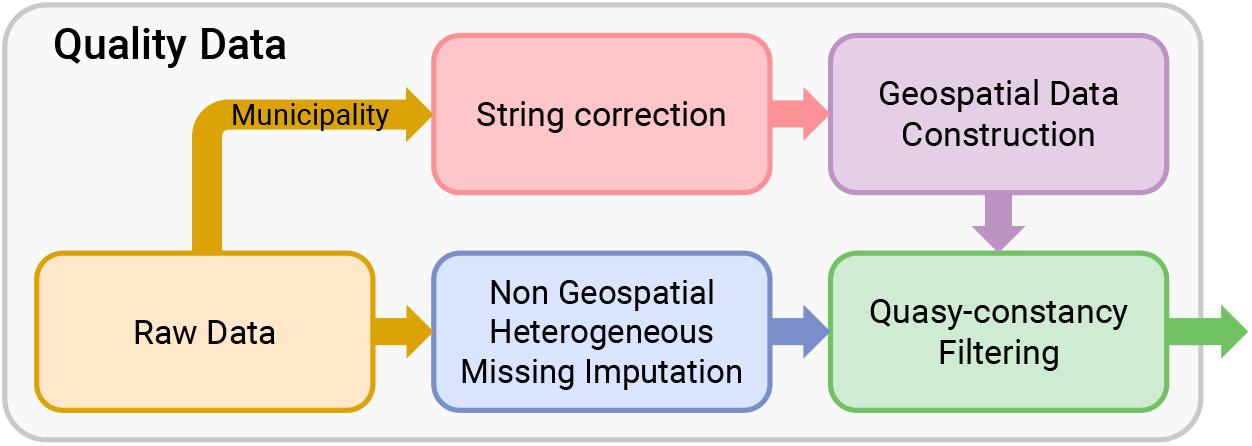
Quality data and information construction process.

#### 2.1.1. Missing values imputation

The quantity of missing values found in the dataset varies significantly among variables. Missing-data imputation consists of replacing missing values in a dataset by appropriate estimated values based on other available information, either in the self-same feature or other measures in the same row. The information used as the estimated value’s explanatory part delimits the imputer model between univariate and multivariate cases[10]. Table 2 shows the different tested imputers.

**Table 2:** Classification of missing values imputers. (^1^)Preserves the original distribution.

|  | Univariate | Multivariate |
| --- | --- | --- |
| **Median**[11] | ✓ |  |
| **Most_frequent**[11] | ✓ |  |
| **Mean**[11] | ✓ |  |
| **A priori probabilistic random**<sup>1</sup> | ✓ |  |
| **KNN imputer (K=5)**[12] |  | ✓ |
| **RF Iterative Imputer**[13][14] |  | ✓ |

Accommodating data to be imputed or deciding between classifier or regressor for the corresponding imputer to be trained requires previously distinguishing between nominal and quantitative variables. Hence, to consider a variable as a nominal variable, a minimum appearance frequency of 5 of every unique value is required, such as shown in [15] for a *χ*^2^ test.

Once the nature of the variable has been established, the nominal explanatory features are hot-encoded internally.

Features are then imputed accordingly to its type. Imputers shown in table 2 are evaluated in order to use the best imputer for each feature. Once the best imputers are selected, features with low imputation score or high missing-value percentage are discarded.

#### 2.1.2. Quasi-constancy filtering

Imbalanced classifiers can be affected by quasi-constant features (variables with very low variability). Applying a quasi-constant filter to delete such variables before classification is an appropriate quality-ensuring step, especially when working with imbalanced data, trying to avoid the tendency of some feature selection models to mistake these kinds of variables as informative variables. For this task, we applied a filter based on Variation [16] and Gini [17] coefficients to look for quasiconstancy by means of a score that ranges from 0 (constancy) to 1 (high variability).

#### 2.1.3. Dataset extension with geospatial data

Taking advantage of the availability of each patient’s municipality record, additional geospatial information was introduced in the dataset, such as pollution [3], population density, or income per capita apart from the original 242 variables. These variables could potentially be related to the T2D disease.

The municipalities column is interesting not by itself but because it links to other types of relevant information. The final municipality corrected strings [8] have been used as the key to link with external datasets related to pollution and family income, allowing the construction of new geospatial information.

On the one hand, the pollution dataset is an aggregated picture of a past moment from [9]. It only shows information for a subset of all the municipalities with geographical sensors, formed by 381 municipalities from a total number of 8112 (*≈* 95% missing values). However, since we know the latitude and longitude of the municipalities, it is possible to approximate missing values by using a function, such is shown in equation 1, which computes the imputed value for the municipality as a weighted mean inverse to the distance squared to the k-nearest municipalities. On the other hand, mean rent by unit of consumption, obtained from [18], shows only *≈* 20% missing values that are imputed by using the same equation 1.

Let *f*_*m*_ be the pollution feature assigned for a missing municipality *m, M* the set of nearest municipalities to the municipality *m* with information, *k* one of the nearest municipalities to the municipality *m* to impute, *d*_*m*_*k* the *L*^2^ distance between the municipality *m* and the municipality *k*, and *N* a normalization factor.

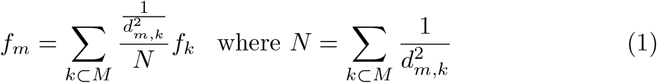

The pollution data, after missing-data imputation, is related to either one of these kinds of **particles**: 1) *Suspended particles <* 2.5*μM*, 2) *Suspended particles <* 10*μM*, **metals**: 1) *Cd(PM10)*, 2) *Ni(PM10)*, 3) *Pb(PM10)*, 4) *As(PM10)*, **gases**: 1) *NO*, 2) *NO*_*2*_, 3) *SO*_*2*_, 4) *CO*, **organic molecules**: 1) *Benzo(a)pyrene C*_*20*_*H*_*12*_ *(PM10)*, 2) *BENZENE C*_*6*_*H*_*6*_ *(PM10)*. An example of this imputation can be seen in Figure 2.

**Figure 2:**
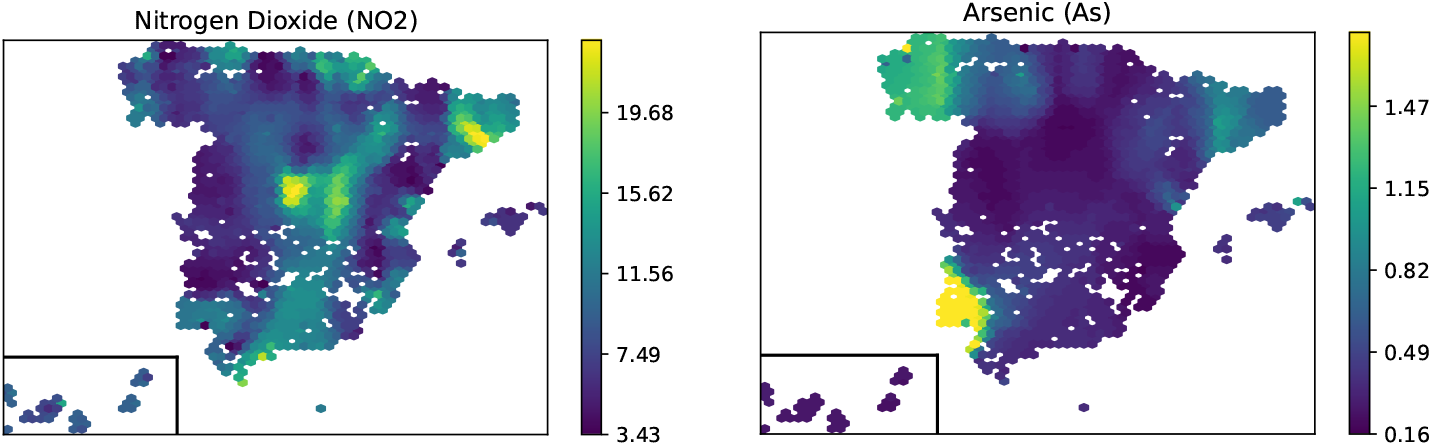
Geographical particle pollution related to *NO*_2_ and *As* respectively. The two different pollution compounds are fully imputed through the available geospatial data interpolation with the formula specified in equation 1.

### 2.2. Model training and evaluation

Four different 10-Fold XGBoost classifier models [19] are fitted, each one focusing on a different role with different economic cost for T2D prediction and diagnosis. Each proposed model selects its features from the most relevant through a permutation-based featured model for AUC-ROC performance [20] with 100 repetitions using a XGBoost-Classifier as the prediction model. The resulting training sets include those features with a left 95% confidence interval for AUC-ROC variation greater than 0.001. Afterwards, with the chosen set of features for each one of the models, the corresponding XGBoost classifier model is trained and evaluated using a 10-fold cross-validation. Finally, prognosis models explainability has been tackled by using Partial dependence estimations [21]. These models can help to identify the factors that can be relevant in the risk of developing T2D, and the differences between patients and healthy people in their behavior and other risk factors. Differences solely between patients can also be assessed to study possible differences between those that know or do not know that they have T2D. In addition, using the clinical parameters used for T2D diagnosis allows us to verify the procedure’s viability.

Two scenarios are proposed:

- The *environmental and lifestyle scenario* has the lowest economic cost because it only requires modifiable and unmodifiable environmental factors to compute the final T2D risk without any biochemistry values and can be obtained directly by the patient with minimal aid.
- The *healthcare scenario* uses a set of general biochemistry values joined to other environmental features to establish the corresponding T2D risk and can be considered of medium economic cost.

For each of these two scenarios, two models are fitted: one for diagnosis (the patient has the T2D at the moment but does not know it yet) and prognosis (each individual could develop T2D up to 7 years after the test but does not present the illness at the moment of data adquisition).

For the creation of each one of the predictive models, individuals can or cannot have T2D. However, all the patients included in the dataset either do not have T2D or they do but still do not know it. The latter group, which are assigned the T2D-positive target, are diagnosed for the first time during data acquisition for this study. This ensures no biases are included due to the lifestyle and treatment adjustments that can be expected once a patient is diagnosed. Besides, in prognosis models all the individuals with T2D presence in the present time are discarded looking for a real prognosis of the illness.

## 3. Results

Firstly, variables with a 90% of missing data have been filtered. Table 3 shows the set of variables filtered by this subject.

**Table 3:** Variables filtered by high missing values proportion (*τ >* 90%).

| Features | Missing values (%) |
| --- | --- |
| Second psychotropic (PSIC_CL2) | 95.71% |
| Second analgesic (AINE_CL2) | 99.26% |
| Second hypotensive (HIPO_CL2) | 90.77% |
| Third hypotensive (HIPO_CL3) | 97.21% |
| Fourth hypotensive (HIPO_CL4) | 99.42% |

Next, the remaining variables with missing values are imputed by the best individual corresponding imputer model. The subset of variables that do not reach a minimum score of 70% after imputation is also filtered out as it is commented previously in section 2.1.1. The quasi-constancy variable filter is the last data quality step to be applied. Table 4 shows the variables filtered by lower missing value imputer model performance and the ones filtered out due to quasi-constancy.

**Table 4:** Variables filtered by lower missing value imputer accuracy (*<* 70%) and quasiconstancy.

| Imputation quality filtering | Quasi-constancy filtering |
| --- | --- |
| Menopause treatment | Pregnant |
| Gestational diabetes | Taking alpha-blockers |
| Causes amenorrhoea | Serious illness |
| Other infusions | Taking uricosorics |
| Physic exercise type | Number of children with T2D |
| Disease | Peripheral vasculopathy |
| Analgesic (AINES_CL) | Lead pollution |
| First psychotropic (PSIC_CL1) | Surgery |
|  | Surgery |
| | BMI $\geq 40$ |
| | BMI $\leq 19$ |
|  | Birth less than 6 months ago |
|  | Taking renin inhibitors |

Finally, four different 10-Fold XGBoost models are fitted. The two evaluated scenarios, named *environmental and lifestyle* and *healthcare*, are described in Section 2.2. The required features and performances of the diagnosis models for each of the two scenarios are shown in Figure 3 and Figure 4 respectively. Table 5 describes the final environmental chosen features for each one of the different models.

**Table 5:** Environmental features description used for the different models. Models: 1 Environmental and lifestyle, 2 Healthcare.

| Features | Models |
| --- | --- |
| Age | 1,2 |
| Waist to Hip Ratio (WHR) | 1,2 |
| Waist | 1,2 |
| # Parents with T2D | 1 |
| Body Mass Index (BMI) | 1,2 |
| Chocolate | 1 |
| Physical activity | 1 |
| Sugar, honey, candies, jam | 1 |
| Non-sugar soft-drinks number | 1 |
| # Brothers with T2D | 1 |
| # Hypotensive drugs (N_HIPOTE) | 1 |
| Civil status | 1,2 |
| Moderate MET-minutes/week (MET) | 1,2 |
| Weight at 18 | 1 |
| Province | 1 |
| Arterial hypertension 90/140 (HTA140) | 1 |
| Benzoapyrene (C<sub>20</sub>H<sub>12</sub>) | 1 |
| # Sugar Soft-drinks | 1 |
| Sweet, cakes, buns | 1 |
| # Hours sleeping | 1 |
| # of times eating outside | 1 |
| # MET-minutes/day | 1 |
| DM family background (AF_DM) | 1 |
| Whole grains | 1 |
| Canned fish, sellfish | 1 |
| Salads | 1 |
| How many cigarretes? | 1 |
| Hematocrit according to patient (HCT) | 1 |
| Basal glucose (mg/dL) (GLUCOSE) | 2 |
| Arsenic (As) | 2 |
| Autonomous community | 2 |
| # Years taking the contraceptive | 2 |
| # Hours sit during the last 7 days | 2 |

**Figure 3:**
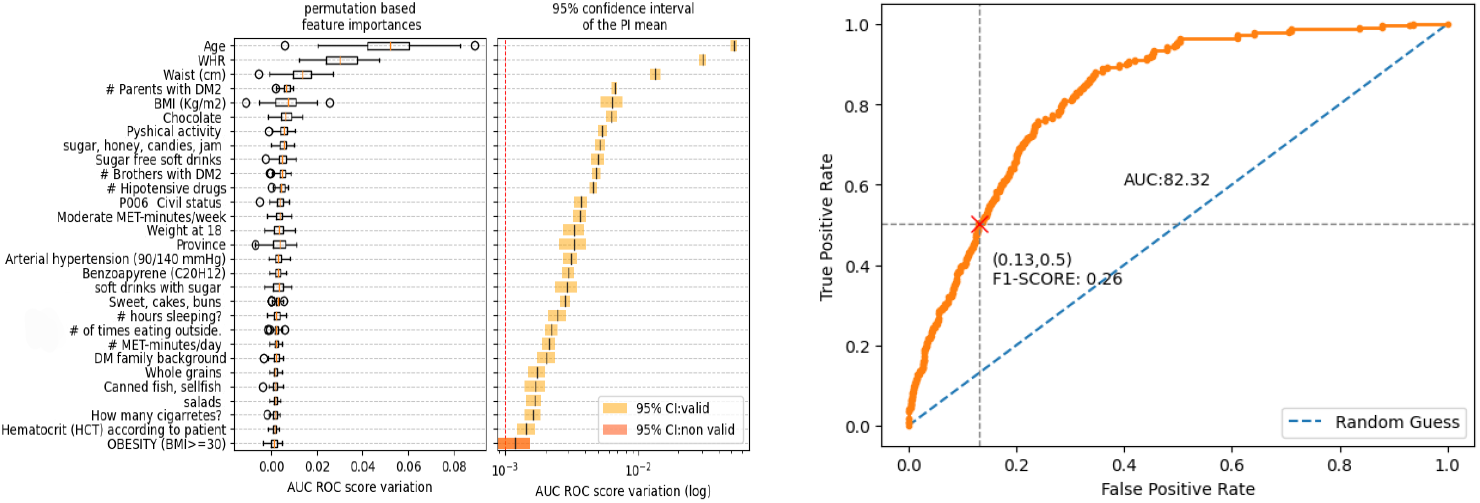
Environmental and lifestyle AUC variable importance for the diagnostic model (left) and 10-fold cross validation ROC curve (right).

**Figure 4:**
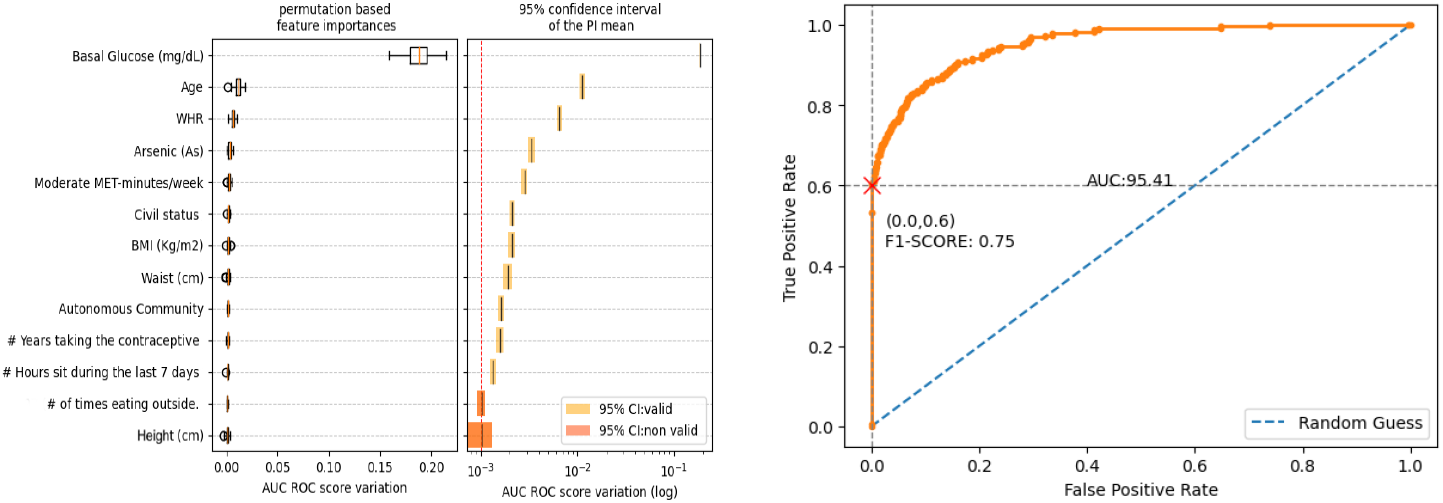
Healthcare AUC variable importance for the diagnostic model (left) and 10-fold cross validation ROC curve (right).

Next, figure 5 shows us the T2D risk prediction for the *environmental and lifestyle* prognosis model trained with individuals without follow-up and tested with individuals with follow-up that do not have T2D in the present but could have T2D or not in a future revision. In summary, results shown in figures 3 and 4 are for T2D diagnostic but the results shown in 5 are for T2D *environmental and lifestyle* prognosis model. Figure 6 show us T2D probability contribution for each explanatory variable used in this latter prognosis model.

**Figure 5:**
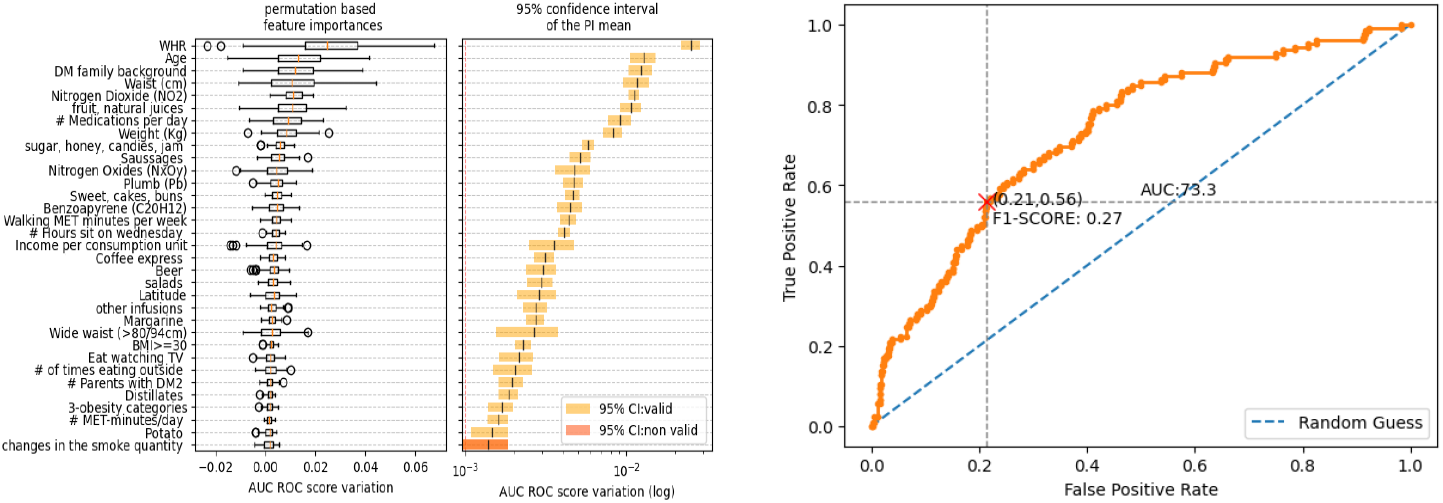
Environmental and lifestyle AUC variable importance for the prognosis model (left) and 10-fold cross validation ROC curve (right).

**Figure 6:**
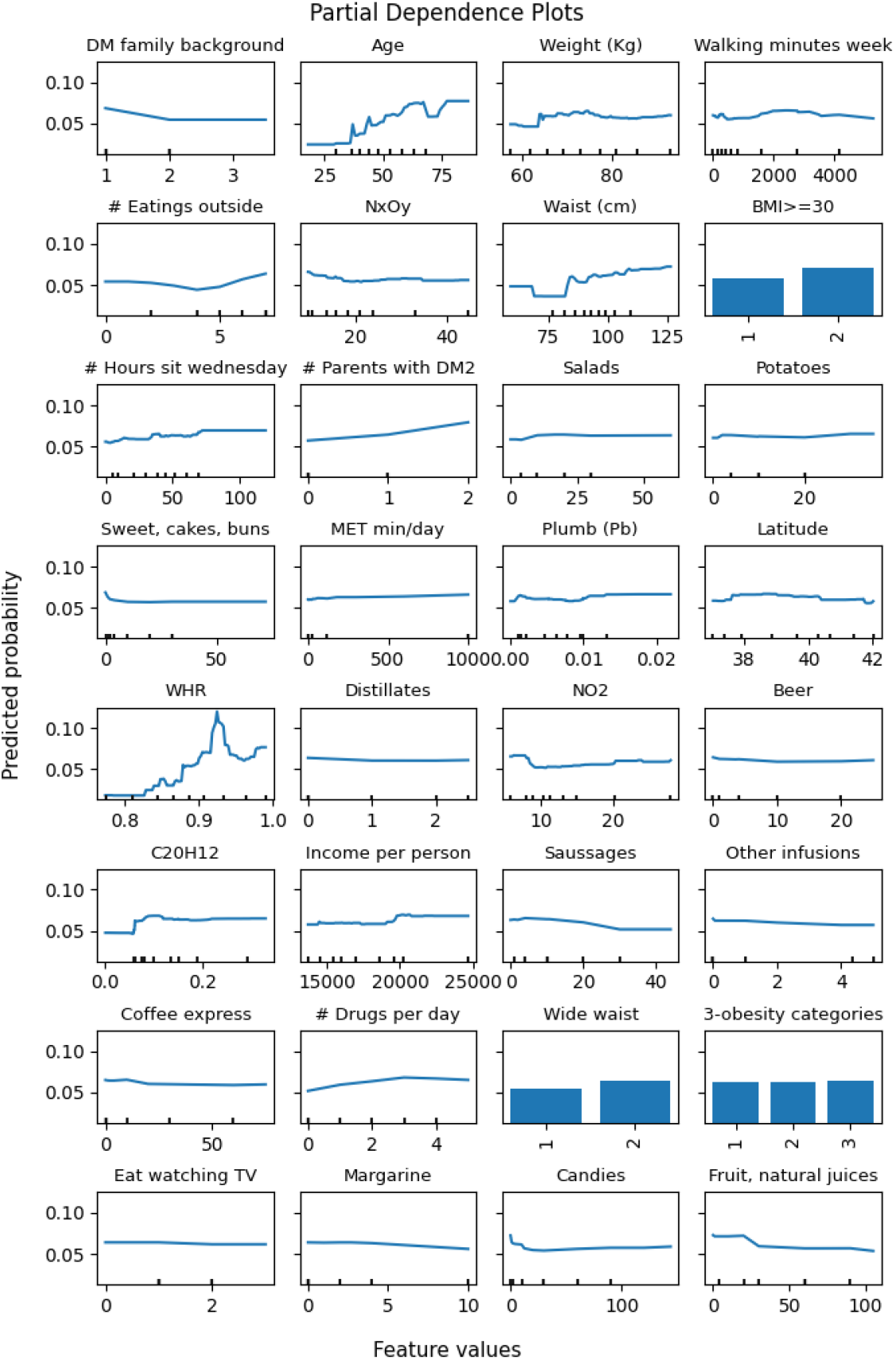
Variable contribution to T2D probability in *Environmental and lifestyle* prognosis model.

Lastly, figure 7 shows us the T2D risk prediction for the *healthcare* prognosis model trained with individuals without follow-up and tested with individuals with follow-up that do not have T2D in the present but could have T2D or not in a future revision. The T2D probability contribution for each explanatory variable used in this *healthcare* prognosis model is presented in Figure 8.

**Figure 7:**
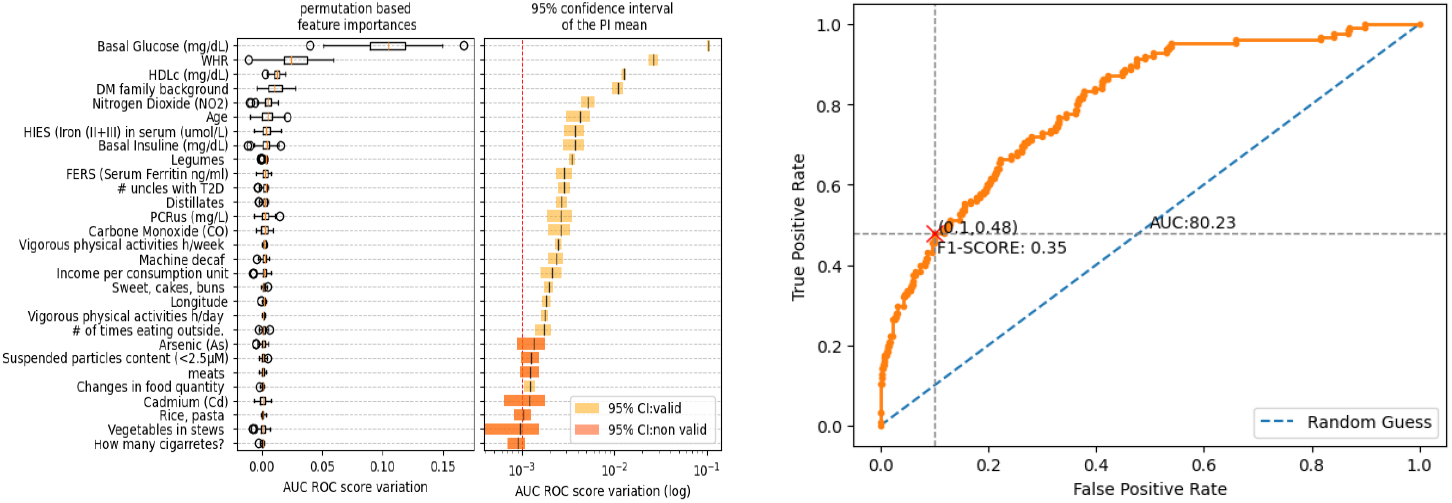
Healthcare AUC variable importance for the prognosis model (left) and 10-fold cross validation ROC curve (right).

**Figure 8:**
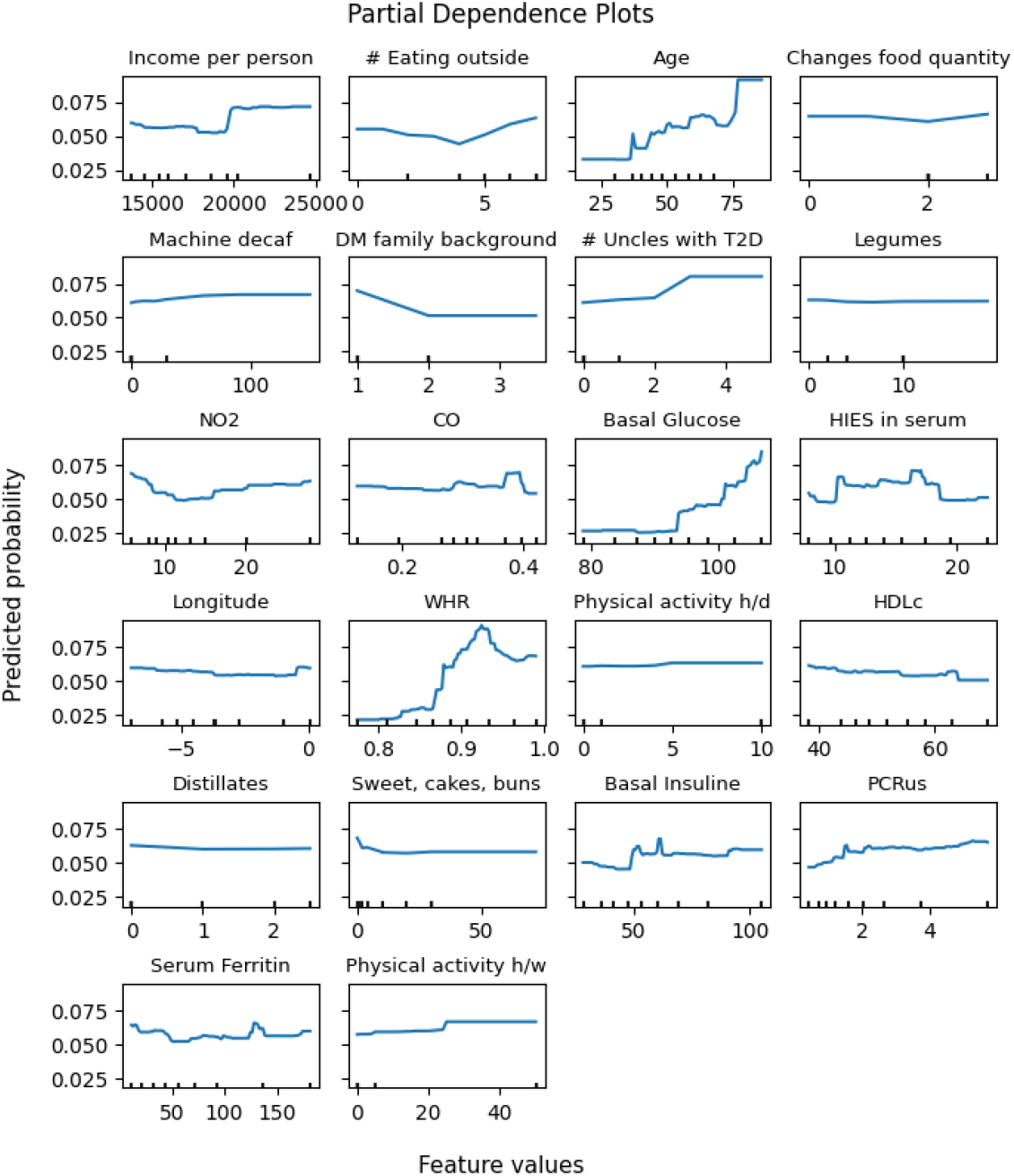
Variable contribution to T2D probability in *Healthcare* prognosis model.

## 4. Discussion

T2D is a complex disease influenced by diverse heterogeneous factors, from genomics to environmental features. Genomics is hoped to be most relevant in future T2D risk detection and disease prediction. Environmental and lifestyle features not only show a relevant role to detect the T2D presence in a patient but also for risk of developing T2D in the future.

The work developed in this article aims to find a way to discover relevant factors related to T2D present in the different heterogeneous environmental features dataset, which requires working with nominal, ordinal and quantitative mixed information and solving different issues present in data. First, the geographical pollution and rent data construction from the municipality field. Secondly, a hybrid missing-value imputation, which allows accommodating a different imputation model for each one of the features in the dataset automatically and independently. Thirdly, a quasi-constancy detection for heterogeneous variables through a new proposed adimensional 0-1 heuristic, which allows us to detect and filter it in an easy and ordered way.

This work proposes the creation of four different models to predict T2D (diagnosis) and also risk of future T2D (prognosis) after using all the tools mentioned above to improve data quality, constructing new features from the available data from heterogeneous sources. Each of the proposed models is intended to be used in different use case scenarios, designed with different roles and different expected costs and performances.

For the record, an additional, more specific model was trained, one that requires a set of more specific biochemistry values joined to some environmental features, as well as the most expensive tests. This model proved to be the best perfomant model, with an almost perfect 99.0 AUC-ROC. This suggests that current clinical tests are accurate enough in T2D diagnosis. The explanatory features used for this classifier are: GLUCOSE, OGTT GLUCOSE and CAPILLARY OGTT GLYCEMIA.

The first model, aimed at identifying patients at risk of suffering from T2D, indicates that many different factors are involved or related to T2D presence. Many of them are known as anthropometric factors (WHR, waist, BMI), familiar history or genetic background (Parents or brothers with T2D), and behavior (physical activity, eating, hours of sleep). Other remarkable aspects include hypotensive drugs, civil status, province, contaminants (benzopyrene), etc. The second scenario, *healthcare*, requires fewer variables as we incorporate the parameters used for diagnosis. In this way, it is interesting that arsenic, civil status, behavior, and anthropometric variables are important in the model, although glucose levels are the main factor, as could be expected.

In the same way, the prediction model for disease development without biochemical data includes many factors affecting the risk as anthropometric (WHR, waist, weight), age, family history, contaminants (NO2, NxOy, Pb, benzopyrene), behavior (eating, walking, hours sitting, coffee, beer, etc.). In the last model, the prediction of T2D using biochemical parameters, the main factor is the basal glucose levels. Many other parameters are included here, such as anthropometric (WHR), biochemical parameters (basal insulin, HLD, HIES, FERS, PCR), family background, age, contaminants (NO2, CO, As, Cd), and behavior (eating, drinking, exercise). It is clear that many factors affect T2D, and our results highlight the importance of factors that often are not included as risk factors, such as urban contamination.

The presented diagnosis models for *environmental and lifestyle* and *healthcare* scenarios, aimed for a broader, more general population, without needing very specific tests, achieve performances of 82.32 and 95.41 AUC-ROC respectively. On the other hand, prognosis models perform slightly worse than diagnosis, with 73.3 and 80.23 AUC-ROC respectively. Their good performance suggest these models could aid clinicians during diagnosis and prognosis. It is worth noting that the difference in performances for the same kind of models are directly related to the kind of explanatory variables used for the model and their economic cost.

## 5. Conclusions

Different models for diagnosis and prognosis of T2D have been designed, each using different subsets of data preprocessed with the previously mentioned tools. The created models obey three different role-based use cases, observing an increasing economic cost as the model performance also increases, as more specialized biochemistry features become available to assess the presence of the illness. It is essential to observe that the performance of the specialist model reaches a quasi-perfect level, which also informs us about the actual illness detection power we already have by using the well-known specific biochemistry tests related to T2D. Furthermore, it is relevant to remark that the cost of the model is linked to the kind of specific tests required for each one of the models. The lowest cost is achieved by the *environmental and lifestyle* model that only requires that the patient completes a survey with a minimal level of guidance required. Alternatively, new *environmental and lifestyle* and *healhcare* models have been obtained as prognosis models for estimating the risk of developing T2D in the future years. Finally, the feature contributions to T2D prediction for both models in both scenarios have been shown.

## 6. Future Work

Generally, classifier models take a set of parameters and return some probability or risk score on each target class, which can be summarized in a single, expected class prediction. However, other approaches are also viable. Instead of assigning a binary class straight away, we could estimate to which risk percentile a single patient pertains by comparing their prediction with those of a significant, random general population sample. While percentiles can be hard to understand, bounded ranges of percentiles could be converted to a reasonable set of risk categories, such as low, moderate or high risk, imitating a traffic-light system. Being classified into a high-risk tier could prompt the patient to seek further professional help or consider adopting a healthier lifestyle.

Nevertheless, some aspects like age, genetics, ancestry or the place of residence cannot be changed to lower one’s risk of developing the disease. Comparing patients with very different unmodifiable characteristics could be misleading as to how well someone’s lifestyle is doing to prevent the disease. Thus, two different risk scores seem to be interesting. First, comparing the general population would stratify patients by their actual risk. Then, on the other hand, a separate comparison with patients with similar unchangeable characteristics would make a fairer evaluation of someone’s risk factors associated with their lifestyle only, which the patient could try to modify to lower their level of risk.

A new line related to future T2D risk prediction is intended to be carried out by mixing environmental and lifestyle features with genomic data by selecting single-nucleotide polymorphism (SNPs) that appear to be related in any way to T2D disease. Further studies in this direction could take profit of genomic SNPs feature selection done in a previous work during the last years by applying an innovative ensemble feature selection algorithm explained in [22]. The final result of the algorithm was an ordered list of the most voted features that are directed related to feature relevance concerning T2D, which allowed biologists to reduce cost during measuring part, focusing only on the most relevant features for new individuals study.

## Data Availability

The dataset used in the present study is available upon reasonable request to the authors of the Study.

## 7. Acknowledgements

The activities described in this document are part of the Cervera Network of Excellence in Data-Based Enabling Technologies (AI4ES) project, which is cofinanced by the Center for Industrial Technological Development, E.P.E. (CDTI) and by the European Union through the Next Generation EU Fund, within the Cervera aid program for Technology Centers by the CDTI, with file number CER-20211030.

Research grants PI17/00544 and PI21/00506 from Ministerio de Ciencia e Innovación and Instituto de Salud Carlos III (ISCIII). CIBERDEM (CB07/08/0018) is part of CIBER (Consorcio Centro de Investigación Biomédica en Red) funded by ISCIII. Both research projects and CIBER are co-funded by the European Union (European Regional Development Fund (ERDF) “A way to build Europe”).

The project is a collaborative study with various phases and subprojects in which a large number of researchers and technicians have collaborated, to whom we are indebted. We are especially grateful to the Steering Committee of the study together with all collaborators who have made it possible (https://www.sediabetes.org/cientifico-yasistencial/investigacion/proyectos-deinvestigacion/estudio-dibet-es/, accessed on 17 June 2023). Di@bet.es-incidence Study Group: G. Rojo-Martínez, F. Soriguer, S. Valdés, N. Colomo, C. Maldonado, E. García-Escobar, A. Lago-Sampedro and S. García-Serrano (Centro de Investigación Biomédica en Red de Diabetes y Enfermedades Metabólicas Asociadas [CIBERDEM]; UGC Endocrinología y Nutrición, Hospital Regional Universitario, Instituto de Investigación Biomédica de Málaga [IBIMA], Malaga, Spain), A. Goday (Servicio de Endocrinología y Nutrición, Hospital del Mar, Barcelona, Spain), E. Bordiú (Laboratorio de Bioquímica, Hospital Universitario San Carlos, Madrid, Spain), A. Calle-Pascual (Servicio de Endocrinología y Nutrición, Hospital Universitario San Carlos, Madrid, Spain and CIBERDEM), L. Castaño and I. Urrutia (CIBERDEM; Unidad de Investigación, Hospital Universitario Cruces, Universidad del País Vasco/Euskal Herriko Unibertsitatea [UPV/EHU], Baracaldo, Vizcaya, Spain), C. Castell (Servicio de Prevención enfermedades crónicas no transmisibles, Departamento de Salud. Barcelona, Spain, E. Delgado and E. Menéndez (Servicio de Endocrinología y Nutrición, Hospital Central de Asturias, Oviedo, Asturias, Spain), J. Franch-Nadal (Equipo de Atención Primaria Raval Sud, Institut Català de la Salut, Red GEDAPS [Grupo de Estudio de la Diabetes en Atención Primaria de la Salud], Unitat de Suport a la Recerca, Institut d’Investigació en Atenció Primària Jordi Gol, Barcelona, Spain), S. Gaztambide (CIBERDEM; Servicio de Endocrinología y Nutrición, Hospital Universitario de Cruces, UPV/EHU, Baracaldo Vizcaya, Spain), J. Girbés (Unidad de Diabetes, Hospital Arnau de Vilanova, Valencia, Spain), R. Gomis (CIBERDEM; IDIBAPS, Hospital Clínic de Barcelona, Barcelona, Spain), E. Montanya (Bellvitge Hospital-IDIBELL, University of Barcelona, Hospitalet de Llobregat, Barcelona, Spain), E. Ortega (CIBEROBN; IDIBAPS, Hospital Clínic de Barcelona, Barcelona, Spain), I. Ramis (CIBERDEM, Spain), J. Vendrell (CIBERDEM; Rovira i Virgili University; Servicio de Endocrinología y Nutrición, Hospital Universitario Joan XXIII, Institut d’Investigacions Sanitàries Pere Virgili, Tarragona, Spain).

## References

[1] J. Xie, M. Wang, Z. Long, H. Ning, J. Li, Y. Cao, Y. Liao, G. Liu, F. Wang, A. Pan, Global burden of type 2 diabetes in adolescents and young adults, 1990-2019: systematic analysis of the global burden of disease study 2019, BMJ 379 (2022).

[2] Global obesity observatory, https://data.worldobesity.org/tables/prevalence-of-adult-overweight-obesity-2/, 2023. Accessed: 2023-01-10.

[3] Y. Li, L. Xu, Z. Shan, W. Teng, C. Han, Association between air pollution and type 2 diabetes: an updated review of the literature, Therapeutic Advances in Endocrinology and Metabolism 10 (2019) 2042018819897046. PMID: 31903180.

[4] R. S. Surwit, M. S. Schneider, M. N. Feinglos, Stress and diabetes mellitus, Diabetes Care 15 (1992) 1413–1422.

[5] F. A. Wenzl, S. Kraler, G. Ambler, C. Weston, S. A. Herzog, L. Räber, O. Muller, G. G. Camici, M. Roffi, H. Rickli, K. A. A. Fox, M. de Belder, D. Radovanovic, J. Deanfield, T. F. Lüscher, “sex-specific evaluation and redevelopment of the grace score in non-st-segment elevation acute coronary syndromes in populations from the uk and switzerland: a multinational analysis with external cohort validation”, The Lancet 400 (2022) 744–756.

[6] H. W. H., The economics of diabetes prevention, The Medical clinics of North America 95 (2011) 373–viii.

[7] F. Soriguer, A. Goday, A. Bosch-Comas, E. Bordiú, A. Calle-Pascual, R. Carmena, R. Casamitjana, L. Castaño, C. Castell, M. Catalá, E. Delgado, J. Franch, S. Gaztambide, J. Girbés, R. Gomis, G. Gutiérrez, A. López-Alba, M. T. Martínez-Larrad, E. Menéndez, I. Mora-Peces, E. Ortega, G. Pascual-Manich, G. Rojo-Martínez, M. Serrano-Rios, S. Valdés, J. A. Vázquez, J. Vendrell, Prevalence of diabetes mellitus and impaired glucose regulation in spain: the study, Diabetologia 55 (2012) 88–93.

[8] J. R. Navarro-Cerdan, R. Llobet, J. Arlandis, J.-C. Perez-Cortes, Composition of constraint, hypothesis and error models to improve interaction in human–machine interfaces, Information Fusion 29 (2016) 1–13.

[9] M. para la Transición Ecológica y el Reto Demográfico, Datos oficiales calidad del aire 2021, https://www.miteco.gob.es/es/calidad-y-evaluacion-ambiental/temas/atmosfera-y-calidad-del-aire/calidad-del-aire/evaluacion-datos/datos/Datos_oficiales_2021.aspx, 2021. [Online; accessed 20-February-2023].

[10] R. Lall, How multiple imputation makes a difference, Political Analysis 24 (2017) 414–433.

[11] Simpleimputer, https://scikit-learn.org/stable/modules/generated/sklearn.impute.SimpleImputer.html, 2023. Accessed: 2023-02-15.

[12] O. Troyanskaya, M. Cantor, G. Sherlock, P. Brown, T. Hastie, R. Tibshirani, D. Botstein, R. B. Altman, Missing value estimation methods for DNA microarrays, Bioinformatics 17 (2001) 520–525.

[13] S. F. Buck, A method of estimation of missing values in multivariate data suitable for use with an electronic computer, Journal of the Royal Statistical Society. Series B (Methodological) 22 (1960) 302–306.

[14] S. van Buuren, K. Groothuis-Oudshoorn, mice: Multivariate imputation by chained equations in r, Journal of Statistical Software 45 (2011) 1–67.

[15] D. S. Yates, D. S. Starnes, D. S. Moore, The practice of statistics, W H Freeman & Co, 2002.

[16] L. H. Koopmans, D. B. Owen, J. I. Rosenblatt, Confidence intervals for the coefficient of variation for the normal and log normal distributions, Biometrika 51 (1964) 25–32.

[17] G. M. Giorgi, C. Gigliarano, The gini concentration index: A review of the inference literature, Journal of Economic Surveys 31 (2017) 1130–1148.

[18] I. N. de Estadística, Atlas de distribución de renta de los hogares, https://ine.es/dynt3/inebase/index.htm?padre=7132, 2023. [Online; accessed 20-February-2023].

[19] T. Chen, C. Guestrin, XGBoost: A scalable tree boosting system, in: Proceedings of the 22nd ACM SIGKDD International Conference on Knowledge Discovery and Data Mining, KDD ‘16, ACM, New York, NY, USA, 2016, pp. 785–794. URL: http://doi.acm.org/10.1145/2939672.2939785. doi:10.1145/2939672.2939785.

[20] Permutation feature importance, https://scikit-learn.org/stable/modules/permutation_importance.html, 2023. Accessed: 2023-06-01.

[21] Partial depence, https://scikit-learn.org/stable/modules/generated/sklearn.inspection.partial_dependence.html, 2023. Accessed: 2023-06-01.

[22] F. Signol, L. Arnal, J. R. Navarro-Cerdán, R. Llobet, J. Arlandis, J.-C. Perez-Cortes, Seqens: An ensemble method for relevant gene identification in microarray data, Computers in Biology and Medicine 152 (2023) 106413.

